# Novel AI-Driven Infant Meningitis Screening from High Resolution Ultrasound Imaging

**DOI:** 10.1101/2024.08.29.24312709

**Authors:** Hassan Sial, Francesc Carandell, Sara Ajanovic, Javier Jiménez, Rita Quesada, Fabião Santos, W. Chris Buck, Muhammad Sidat, UNITED Study Consortium, Quique Bassat, Beatrice Jobst, Paula Petrone

**Affiliations:** Biomedical Data Science Team, Barcelona Institute for Global Health (ISGlobal), Barcelona, Spain; Kriba, Barcelona Science Park, Barcelona, Spain; Barcelona Institute for Global Health (ISGlobal) - Hospital Clínic, Universitat de Barcelona, Barcelona, Spain; University of California Los Angeles David Geffen School of Medicine, Los Angeles, USA; Universidade Eduardo Mondlane Faculdade de Medicina, Maputo, Mozambique; Centro de Investigação em Saúde de Manhiça (CISM), Maputo, Mozambique; ICREA, Pg. Lluís Companys 23, Barcelona, Spain; Pediatrics Department, Hospital Sant Joan de Déu, Universitat de Barcelona, Barcelona, Spain; CIBER de Epidemiología y Salud Pública, Instituto de Salud Carlos III, Madrid, Spain

**Author notes:** **Address for correspondence:** Paula Petrone, Associate Research Professor, Barcelona Institute for Global Health (ISGlobal), C/ Doctor Aiguader, 88. 08003. Barcelona, Spain. Contributed equally as co-authors: Hassan Sial, Francesc Carandell. Contributed equally as co-senior authors: Beatrice Jobst, Paula Petrone. UNITED Study Consortium collaborators are listed in the acknowledgment section.

**Keywords:** Infant Meningitis, Ultrasound Imaging, Deep Learning, Neosonics^®^, Explainable AI, Non-invasive Screening, Lumbar Puncture, White Blood Cell Count

## Abstract

**Background:** Infant meningitis can be a life-threatening disease and requires prompt and accurate diagnosis to prevent severe outcomes or death. Gold-standard diagnosis requires lumbar punctures (LP), to obtain and analyze cerebrospinal fluid (CSF). Despite being standard practice, LPs are invasive, pose risks for the patient and often yield negative results, either because of the contamination with red blood cells derived from the puncture itself, or due to the disease’s relatively low incidence due to the protocolized requirement to do LPs to discard a life-threatening infection in spite its relatively low incidence. Furthermore, in low-income settings, where the incidence is the highest, LPs and CSF exams are rarely feasible, and suspected meningitis cases are generally treated empirically. There’s a growing need for non-invasive, accurate diagnostic methods.

**Methodology:** We developed a three-stage deep learning framework using Neosonics^®^ ultrasound technology for 30 infants with suspected meningitis and a permeable fontanelle, from three Spanish University Hospitals (2021-2023). In Stage 1, 2194 images were processed for quality control using a *vessel/non-vessel* model, with a focus on vessel identification and manual removal of images exhibiting artifacts such as poor coupling and clutter. This refinement process led to a focused cohort comprising 16 patients—6 cases (336 images) and 10 controls (445 images), yielding 781 images for the second stage. The second stage involved the use of a deep learning model to classify images based on WBC count threshold (set at 30 cells/mm^3^) into *control* or *meningitis* categories. The third stage integrated eXplainable Artificial Intelligence (XAI) methods, such as GradCAM visualizations, alongside image statistical analysis, to provide transparency and interpretability of the model’s decision-making process in our AI-driven screening tool.

**Results:** Our approach achieved 96% accuracy in quality control, 93% precision and 92% accuracy in image-level meningitis detection, and 94% overall patient-level accuracy. It identified 6 meningitis cases and 10 controls with 100% sensitivity and 90% specificity, demonstrating only a single misclassification. The use of GradCAM-based explainable AI (XAI) significantly enhanced diagnostic interpretability, and to further refine our insights, we incorporated a statistics-based XAI approach. By analyzing image metrics like entropy and standard deviation, we identified texture variations in the images, attributable to the presence of cells, which improved the interpretability of our diagnostic tool.

**Conclusion:** This study supports the efficacy of a multistage deep learning model for the non-invasive screening of infant meningitis and its potential to guide indications of LPs. It also highlights the transformative potential of AI in medical diagnostic screening for neonatal healthcare and paves the way for future research and innovations.

## 1. Introduction

Infant meningitis, an inflammatory condition of the protective membranes covering the brain and spinal cord, is a life-threatening disease, especially among newborns and infants. Early and precise diagnosis is critical to rapidly initiate treatment, and thus to prevent severe, lifelong neurological *sequelae* and reduce the high mortality rates associated with the disease [1]. However, the diagnostic challenges appear significant, with estimates from 2019 suggesting approximately 2.51 million new cases and 336,000 deaths across all age groups, globally. Meningitis cases disproportionately cluster among young infants and children under five, which account for up to 1.28 million new cases and roughly 112,000 fatalities annually, bearing nearly half of the mortality attributed to meningitis [2].

The current standard diagnostic procedure involves lumbar punctures (LPs) to obtain cerebrospinal fluid (CSF) for the confirmatory laboratory analysis. Confirmation of meningitis needs an elevated white blood cell (WBC) count and/or microbiological confirmation (either through CSF culture or molecular techniques to identify the causative pathogen). LPs are not innocuous and can cause infections, bleeding, nerve damage, or respiratory arrests, particularly among the youngest infants. They also require specialized equipment, skilled personnel, and an accompanying laboratory infrastructure to process and analyze the CSF [3]. The limitations of LP are most acutely felt in resource-poor settings, which often leads to critical delays in diagnosis and subsequent treatment, thereby increasing mortality. In high-income countries (HICs), a proactive stance is often adopted, with LPs performed as part of standard protocols on the youngest infants, perceived to be at a higher risk of disease, to prevent any missed diagnoses of Acute bacterial meningitis (ABM), given its high associated lethality. However, given the relatively low incidence of meningitis disease, less than five percent of these punctures yield a positive result [5,6]. Even in well-equipped facilities, complications like the blood contamination of cerebrospinal fluid samples during LP can challenge accurate diagnosis, exposing the patient to preventive, yet often unnecessary antibiotic treatment [4]. These limitations highlight the necessity for an accurate, non-invasive screening approach to screen (i.e., rule in/rule out) meningitis early in infants. Alternatively, this approach may target infants under high suspicion of meningitis, who may require a confirmatory lumbar puncture.

Neonates and young infants, who are at the highest risk of meningitis, possess a unique anatomical feature on their cranium known as the permeable fontanel. This feature, due to the immature ossification of the cranium until approximately the first year of life, provides a distinct advantage for visualizing the CSF space beneath using ultrasound. The acoustic properties of the fontanel differ significantly from the ossified bones in older children and adults, which ultrasound waves generally cannot penetrate later in life. Leveraging this anatomical particularity in this vulnerable age group, we have developed our technology.

Artificial Intelligence (AI), especially through deep learning, has revolutionized medical diagnostics, including ultrasound imaging. This technology enhances diagnostic precision in a range of conditions and sometimes is on par with human expert analysis [7]. At the heart of this innovation are convolutional neural networks (CNNs), which have become a fundamental element in image analysis. Renowned CNN architectures like AlexNet, VGG, Inception, MobileNet, and ResNet have demonstrated their value in various diagnostic applications [8–17]. The combination of AI with ultrasound is particularly promising for accurate diagnosis in diverse medical conditions, including respiratory diseases such as COVID-19 and pneumonia [18,19], as well as for identifying and classifying tumors, lesions [20,21], and nodules [22].

Our study builds on the emerging and highly innovative research into non-invasive ultrasound methods for potential meningitis screening [23,24], initially focusing on in vitro data. While ultrasound is inherently non-invasive, it can be prone to noise and clutter, typically positioning it more as a screening tool rather than a definitive diagnostic method. However, integrating AI, specifically deep learning, can significantly amplify its screening capabilities. Prior research has shown the potential of high-frequency ultrasound images to measure the concentration of diagnostically relevant particles and cells in laboratory samples [23,25,26]. In our research, we utilize deep learning to refine the accuracy and reliability of diagnostic meningitis screening using high-resolution (HR) ultrasound images from the CSF region lying below the infant fontanel [37].

Explainable AI (XAI) is gaining more and more relevance in healthcare-related applications, enhancing AI-driven decision-making transparency, vital for bias detection and model interpretability [33, 34]. It empowers clinicians and patients to understand AI outputs, essential in diagnostics and plays a key role in reducing misdiagnosis risks and promoting ethical AI use [35, 36]. Despite XAI’s growth in medical diagnostics, particularly in ultrasound imaging for areas like tumor analysis, thyroid nodule diagnosis, and lung image classification [38, 39, 40], applying XAI to infant meningitis screening via ultrasound faces unique challenges. While HR ultrasound produces well-defined and visible WBC backscatter signals even in very low WBC concentrations suspensions in the absence of acoustically attenuation or absorption media [23], when applied on CSF below the infant fontanel, the acoustic beam distortion attributed to the fontanelle tissue generates instead a WBC specific signal pattern in the images, which can be difficult to interpret. This complicates the task of identifying these crucial diagnostic indicators and makes obtaining clear and interpretable insights from deep learning models challenging but necessary. Acknowledging this, our study integrates advanced XAI techniques such as Gradient-weighted Class Activation Mapping (Grad-CAM) [27] alongside image statistical analysis, including entropy, standard deviation, and gradients, to address these specific hurdles. This approach aligns with current best practices in AI transparency and is particularly important for our task in infant meningitis screening, where visualizing and understanding the diagnostically critical pattern produced by the WBC backscatter signals in CSF is imperative for enhancing the diagnostic process and patient outcomes.

Our team has previously developed Neosonics^®^, a cutting-edge, non-invasive ultrasound technology designed to detect backscatter signals from white blood cells (WBC) in cerebrospinal fluid (CSF) beneath the infant’s fontanelle [37]. Leveraging deep learning (DL) for image analysis, this technology aims to classify patients based on WBC levels in CSF, offering a rapid, non-invasive screening method for infant meningitis.

Our study introduces a novel deep learning methodology [50] that further corroborates and analyses in more depth the initial proof of concept in a clinical study [37]. A key element of our work is the integration of Explainable AI (XAI) techniques, enhancing the interpretability and transparency of the AI’s decision-making process.

This approach involves a three-stage process specifically designed to classify the presence or absence of white blood cells, optimizing the screening process for infant meningitis. Stage 1 is dedicated to automated yet rigorous image quality control, required to maintain the accuracy of the screening process. Stage 2 uses a deep learning architecture in combination with ensemble learning for binary classification to distinguish patients as either control or meningitis cases, by recognizing the presence of WBCs in CSF, indicators of meningitis. Stage 3 addresses the “black box” nature of deep learning models by integrating the Gradient-weighted Class Activation Mapping (Grad-CAM) algorithm [27] into our framework. This Explainable AI (XAI) technique makes the decision-making process of our models transparent, highlighting the crucial regions within the ultrasound images that are most informative in the classification. To deepen our understanding further, we analyze the differences between control and meningitis images using statistical measures like entropy, standard deviation, and gradients. This not only improves the accuracy but also significantly enhances the interpretability of our models enabling the identification of potential biases.

This structured methodology combines rigorous quality control with a classification stage, and the application of the XAI to offer a comprehensive and transparent solution for meningitis screening. This study highlights the potential use of ultrasound imaging in combination with deep learning in reducing the need for invasive lumbar punctures in neonatal and infant care and providing a solution that is efficient, cost-effective [49], and easy to deploy in remote areas where there is most need.

## 2. Methods

The methodological workflow is divided into three sequential stages: Stage 1 utilizes AI models for automated and precise image quality control, essential for maintaining the accuracy of the screening process. Stage 2 utilizes AI models in conjunction with ensemble learning for binary classification. These models are finely tuned to detect patterns indicative of white blood cells (WBCs) in cerebrospinal fluid (CSF), crucial markers for meningitis diagnosis. Stage 3 is dedicated to explaining the algorithms’ predictions and results by integrating various techniques, such as Grad-CAM [27], with statistical measures like entropy, standard deviation, and gradients (see Figure 1).

**Figure 1.**
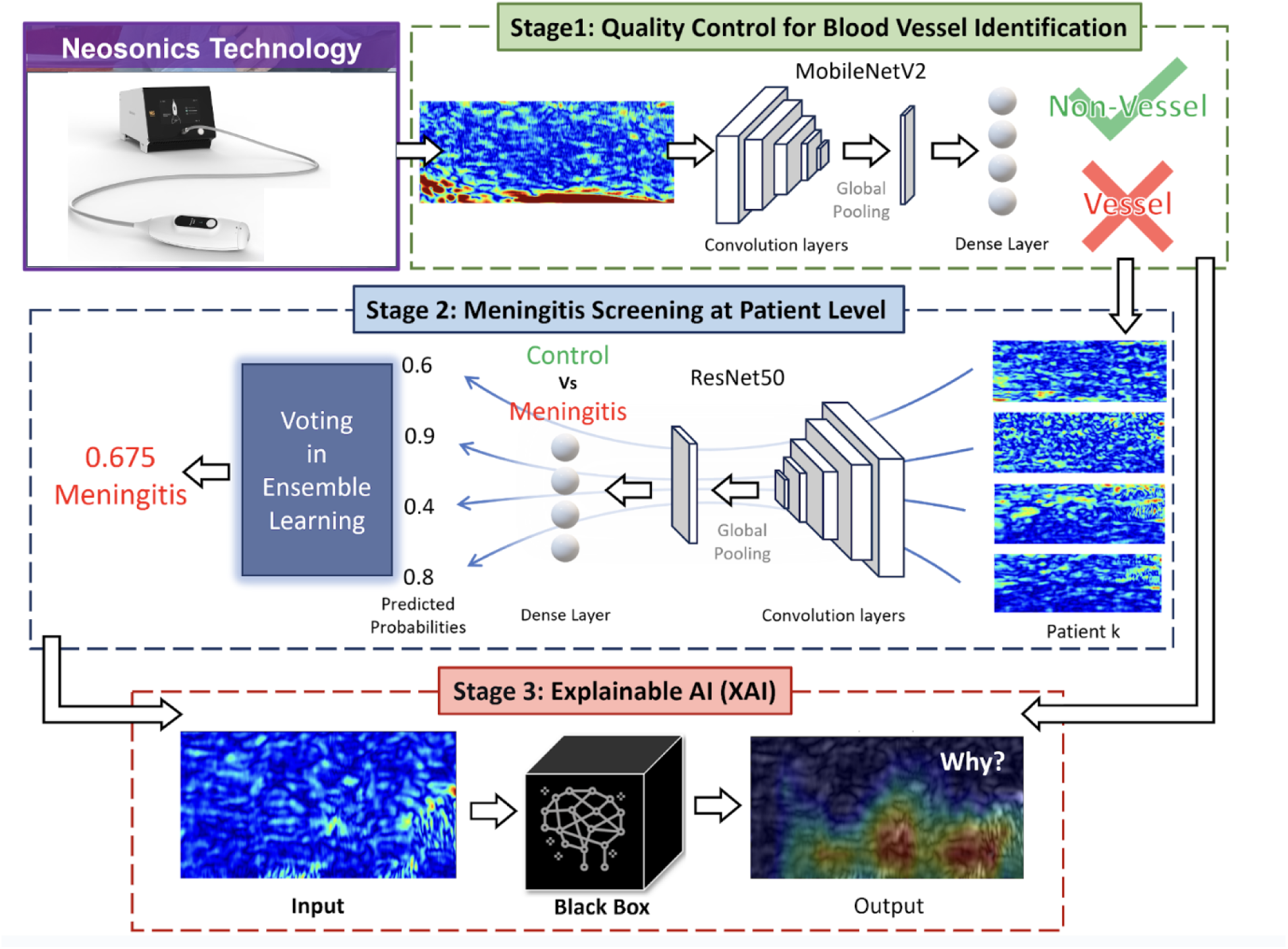
Flowchart of the three-stage methodology employed for infant meningitis diagnostic screening using ultrasound images. Images are acquired on the baby’s fontanelle using novel Neosonics^®^ technology. Stage 1 (Quality Control): MobileNetV2 deep learning architecture is used to filter out ultrasound images that exhibit blood vessels. Stage 2 (Screening) employs binary classification to distinguish between “Control” and “Meningitis” cases based on the presence of increased WBC cellularity in the CSF as visualized in ultrasound images. Stage 3 (Explainable Artificial Intelligence (XAI): XAI techniques are applied for Stage 1 and Stage 2 model interpretability.

### 2.1. Datasets & Ground Truth Generation

**Data acquisition.** A total dataset of 2194 HR ultrasound images was collected from 30 Patients with suspected meningitis, recruited across three hospitals in Spain: La Paz University Hospital, Quirón Salud University Hospital Madrid, and Sant Joan de Déu University Hospital Barcelona [37]. For each patient, a lumbar puncture (LP) was performed within 24 hours before image acquisition for the obtention of cerebrospinal fluid (CSF) samples. Subsequently, HR ultrasound images of the fontanelle region were captured using a specialized device. Clinicians received training in device usage before patient enrollment.

This study rigorously followed ethical guidelines, ensuring patient confidentiality, anonymization of data, and data collection only after obtaining written informed consent from the legal guardians of the infant patients.

**Neosonics technology.** The Neosonics^®^ device is a novel non-commercially available device that has not yet received any certification clearance. It is making spatial scans with steps smaller than 5 microns, which allows capturing the backscatter signals of individual cells within the CSF, crucial for analyzing the composition of serous body fluids in a non-invasive way and at a high sensitivity to structural changes not captured by conventional ultrasound systems [37]. For ultrasound imaging data collection, we used the Neosonics^®^ ultrasound probe positioned over the anterior fontanelle region of the infants’ head. The fontanelle is a spot where the infant’s skull is not yet closed and thus allows the ultrasound signal to pass through to the cerebrospinal fluid lying below the layers of tissue.

**Data preprocessing.** All images were processed to enhance their quality and suitability for analysis, utilizing the signal processing module of the SciPy Python library [46]. The first step involved applying a Butterworth bandpass filter, followed by calculating the signal envelopes using the Hilbert transform and a min-max normalization of the images. The min-max normalization procedure was used to scale the pixel values to a uniform range, maintaining consistency across the dataset.

**Ground Truth Generation.** For Stage 1 (Quality Control), ultrasound images were manually labeled by expert visual inspection to identify the presence or absence of blood vessels in the CSF, as required by the deep-learning model. Of the 2194 high-resolution (HR) ultrasound images reviewed, 965 were labeled as containing blood vessels and 1229 as not containing blood vessels. For visual examples of vessel and non-vessel images, refer to Figure 2-a.

**Figure 2:**
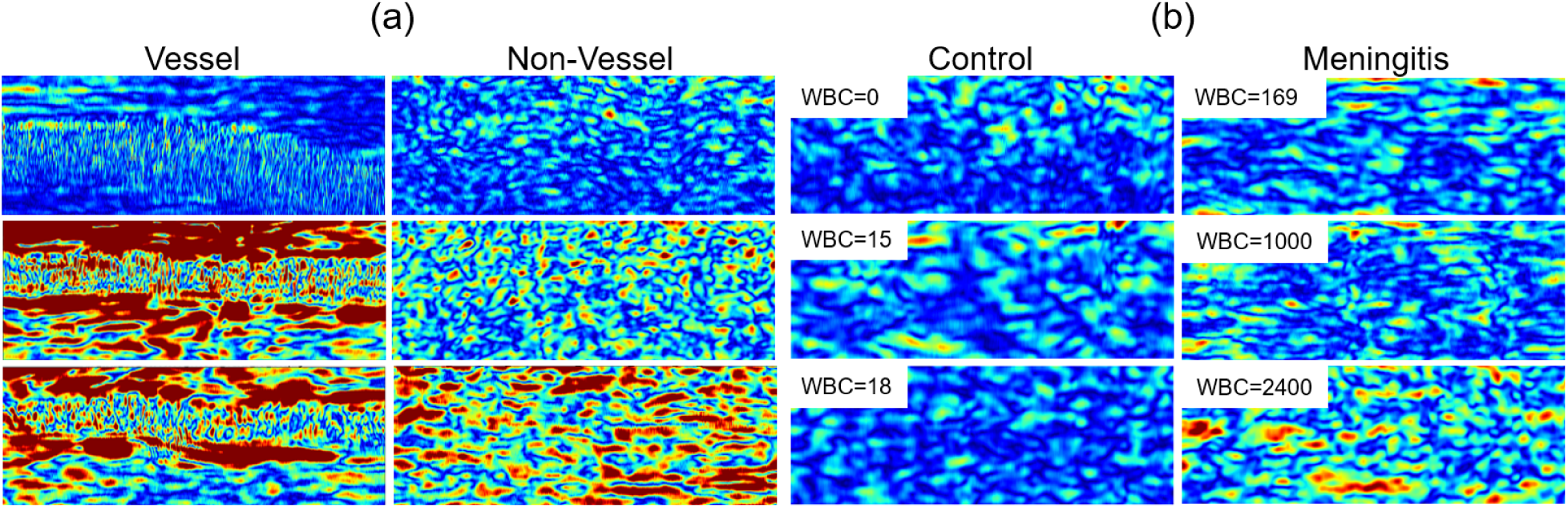
(a) Ultrasound images of the CSF depicting two categories: Presence of vessels vs. no presence of vessels. (b) Ultrasound images of both Control (<30 WBC/mm³) or Meningitis (≥30 WBC/mm³) patients, with the respective WBC count displayed in the top left corner. The colormap used for representing all ultrasound images is jet.

In the screening stage (Stage 2), clinical outcomes derived from WBC counts from LPs were employed to categorize patients as either *control* (<30 WBC/mm³) or *meningitis* (≥30 WBC/mm³), offering a patient-level ground truth. This threshold was selected as a pragmatic criterion to distinguish between likely and unlikely cases of meningitis, in line with clinicians’ consensus and with the anticipation that the majority of Phase I patients would be neonates [37]. Figure 2-b exhibits ultrasound images of patients in both *control and meningitis* categories. Each image also displays the corresponding WBC count in the top left corner.

Following the quality control stage of excluding images with blood vessels present, and additionally excluding images with bad quality or wrong positioning, the cohort was refined to 16 patients (including one patient with 2 LP results), providing a total of 781 HR ultrasound images for Stage 2. (6 cases and 10 controls). On average, there were 40 images acquired per patient. The dataset comprises infants aged 3 to 337 days, with an equal gender distribution (8 males and 8 females) and an average weight of 3,952 grams. Table 1 details a comprehensive summary of the dataset, encompassing demographics, WBC counts, image frame numbers, and clinical outcomes. Further information about this dataset can be found in [37].

**Table 1.** Final Stage Participant Details: CSF: cerebrospinal fluid, LP: lumbar puncture, WBC: white blood cells, WBC in CSF has been reported in units / mm3. The precise ages of the participants have been replaced with quartile-based age ranges. The age ranges were determined by dividing the data into quartiles: 0-9 days, 10-29 days, 30-61 days, and 62+ denoting 62 days and above.

| Participant No. | Age Range (Quartiles, | Weight (grams) | Sex | WBC in CSF * | Number of Images | Clinical final classification | Output of DL algorithm |
| --- | --- | --- | --- | --- | --- | --- | --- |

Table 1. Final Stage Participant Details: CSF: cerebrospinal fluid, LP: lumbar puncture, WBC: white blood cells, WBC in CSF has been reported in units / mm<sup>3</sup>.
| days) |  |  |  |  |  |  |  |
| --- | --- | --- | --- | --- | --- | --- | --- |
| Meningitis |  |  |  |  |  |  |  |
| 1 | 0-9 | 2.035 | f | 53 | 45 | Suspected | Meningitis |
| 2 | 62+ | 5.000 | f | 85 | 24 | Suspected | Meningitis |
| 3 | 0-9 | 3.900 | f | 520 | 32 | Confirmed meningitis | Meningitis |
| 4 | 10-29 | 3.370 | m | 2400 | 50 | Confirmed meningitis | Meningitis |
| 5 | 62+ | 4.900 | m | 300 | 25 | Confirmed meningitis | Meningitis |
| 6 | 30-61 | 3.600 | m | 430 | 36 | Confirmed meningitis | Meningitis |
| 6-2 | 30-61 | 3.600 | m | 169 | 233 | Confirmed meningitis | Meningitis |
| Control |  |  |  |  |  |  |  |
| 7 | 62+ | 4.420 | f | 7 | 11 | No meningitis | Control |
| 8 | 0-9 | 3.290 | f | 3 | 9 | No meningitis | Meningitis |
| 9 | 10-29 | 3.000 | m | 5 | 26 | No meningitis | Control |
| 10 | 10-29 | 2.965 | m | 15 | 14 | No meningitis | Control |
| 11 | 0-9 | 3.890 | f | 5 | 24 | No meningitis | Control |
| 12 | 0-9 | 2.690 | f | 9 | 34 | No meningitis | Control |
| 13 | 30-61 | 4.000 | m | 18 | 81 | No meningitis | Control |
| 14 | 30-61 | 4.150 | m | 0 | 18 | No meningitis | Control |
| 15 | 62+ | 8.600 | m | 11 | 55 | No meningitis | Control |
| 16 | 10-29 | 4.220 | f | 0 | 64 | No meningitis | Control |

### 2.2. Deep Learning Multi Stage Framework

We have developed a deep-learning based framework for both Quality Control in Stage 1 and Meningitis Screening in Stage 2 with an additional focus on Explainable AI in Stage 3 (Figure 1). For all deep learning model development and optimization tasks, we utilized Keras with a TensorFlow backend [47].

#### 2.2.1 Stage 1: Quality Control for Blood Vessel Identification

In this stage, we experimented with two pre-trained models, MobileNetV2 [11] and ResNet50 [12], both initially trained with ImageNet [8], for the classification of *vessel* vs *non-vessel* images. For each model, we added three additional dense layers with units set as (512, 128, 1) on top of the respective pre-trained base models. These added dense layers were the only trainable components, keeping the base model layers frozen during training. This approach allowed us to leverage the advanced feature extraction capabilities of the pre-trained models and tailor them to our specific task of blood vessel identification.

To train and evaluate the models, we employed a 75/25 train/test split combined with a 4-fold cross-validation approach. This setup facilitated a thorough and robust performance evaluation, reducing the likelihood of overfitting and ensuring the generalizability of our findings. A batch size of 16 was used to efficiently process the high-resolution images. The optimization process for both models relied on the Adam optimizer [29], with a learning rate set to 1E-4 to fine-tune the model. Performance was evaluated using a set of standard machine learning performance metrics, including precision, recall, F1 score, and accuracy [28], utilizing the Scikit-learn Python library for this analysis [48].

#### 2.2.2 Stage 2: Meningitis Screening at Patient Level

In Stage 2, a ResNet50 model pre-trained on ImageNet [12] was employed, undergoing fine-tuning to classify individual images as indicative of *meningitis* or *control*. The base layers were kept unchanged, except for the final convolutional block which was adjusted to new data. The network’s fully connected layers were replaced by a global average pooling layer and a single-output dense layer to conduct binary classification.

During training, binary cross-entropy loss is used to distinguish between *meningitis* and *control* cases. We utilized the SGD optimizer [30] for the optimization process, setting a learning rate of 1E-4 to fine-tune the model. Performance was evaluated using a set of standard machine learning performance metrics, including precision, recall, F1 score, and accuracy [28].

For patient-level classification, we adopted a rigorous leave-one-out methodology [28], ensuring that each patient was tested individually while using data from the remaining patients for training. This approach was crucial for the accurate and unbiased evaluation of our model.

Ultrasound images were independently classified, and ensemble-learning techniques were then employed to merge individual image predictions into a unified patient classification. We explored ensemble learning techniques, comparing soft voting with hard voting methodologies [31]. To calculate these, we utilized ‘groupby’ and ‘aggregate’ functions from the Pandas library in Python [41].

The soft voting method involved calculating the average predicted probability score from groups of *N* images for each patient. The process is mathematically expressed as follows:

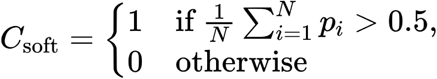

Where *p_i_* is the predicted probability score of meningitis for each image within a group. Scores with an average above 0.5 were classified as *meningitis*, and those below as *control*.

In contrast, the hard voting method was implemented using a majority rule for each group of N images. Each image was initially categorized as *meningitis* or *control* based on its predicted probability score relative to the 0.5 threshold. The final classification for each group was determined by the mode of these binary classifications. This is represented by the following equation:

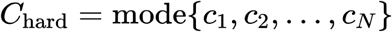

where *c_i_* denotes the class label for each image.

For all experiments within our study, we set *N = 7*.

#### 2.2.3 Stage 3: Explainable AI (XAI)

In this stage, we used Gradient-weighted Class Activation Mapping (GradCAM) [27], using the OmniXAI Python library [43], to gain a deeper understanding of the decision making processes of our convolutional neural networks.

For our analysis, we randomly selected 9 images per patient from both Stage 1 and Stage 2, regardless of their class. These images underwent processing with the GradCAM function, utilizing the final convolutional layer of the deep-learning models from the previous stages. This process generated heatmaps for each image scaled to a 0-255 range with a color spectrum ranging from blue to red. In this spectrum, red areas identified the regions most influential to the network’s classification decisions, while blue areas represented the least influential regions.

We also conducted four statistical comparisons of image characteristics to distinguish *control* from *meningitis* cases of Stage 2. The statistical measures used were: normalized intensity, standard deviation, entropy, and the gradient in the Y direction. These analyses were conducted to identify the backscatter signals of WBCs in case of their presence in the CSF in ultrasound images, yielding a specific pattern in the images of the cerebrospinal fluid, not always easily visible to the human eye. In response to the challenge of visualizing these cells directly, our strategy focused on examining statistical differences between images to uncover patterns that may indicate meningitis.

For normalized intensity, we normalized the pixel intensities of the images to ensure consistency in our calculations. For standard deviation and entropy, we used local standard deviation and local entropy functions from the scikit-image python module [43]. We applied a morphological element in the form of a disk with a size of 5. Finally, for the gradient in the Y direction, we calculated it using the Sobel operator from OpenCV [44].

For each of these measures, we constructed histograms of both *control* and *meningitis* classes to quantitatively analyze the image characteristics. We utilized 20 bins for all histograms, each with an appropriate bin range to capture meaningful variations in the image features. For instance, the normalized intensity histograms ranged from [-1, 1], while the standard deviation histograms spanned [0, 0.2], and entropy histograms covered [0, 8]. These carefully chosen bin sizes and ranges allowed us to extract valuable information from the images. To quantitatively measure the differences between these histograms, we employed the Jensen-Shannon divergence (JSD) [45] method from the SciPy python module [46]. We plotted the relationships of cell count with entropy and mean Grad-CAM score with entropy. For these plots, we fitted a regression line using the linear regression method from the SciPy python module [46] and computed the coefficient of determination (R²) to evaluate the predictive strength and significance of the relationships [28].

## 3. Results

### 3.1 Stage 1: Quality Control

In this initial quality control stage, the primary task was the binary classification of ultrasound images to identify good quality ones and exclude those with blood vessels, crucial for an accurate meningitis screening. Blood vessels can obscure CSF areas, where white blood cells indicate meningitis, potentially leading to screening inaccuracies. We fine-tuned two pre-trained deep learning models, MobileNetV2 [11] and ResNet50 [12]. The performance of these models, including average metrics and their variability as indicated by standard deviation across 4-fold cross-validation, is detailed in Table 2. Examples of vessel/non-vessel images are illustrated in Figure 2-a.

**Table 2.**
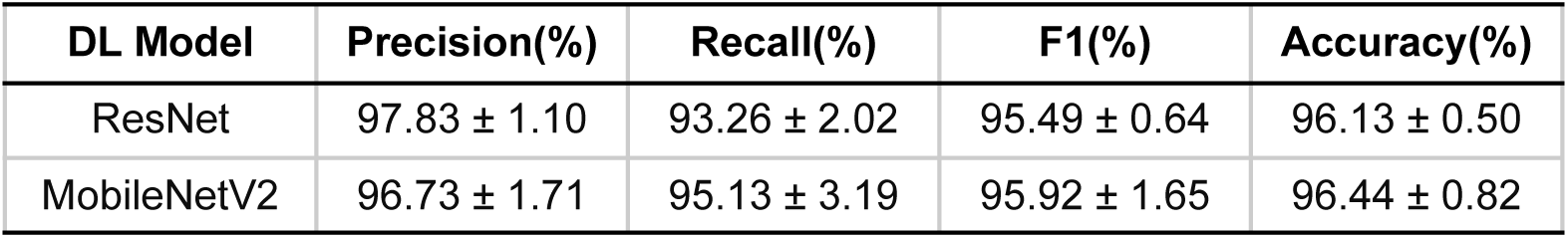
Performance Comparison of MobileNetV2 and ResNet50 Models Using 4-Fold Cross-Validation for Stage 1 (Quality Control), Including Mean Metrics and Standard Deviation to Indicate Variability Across Folds.

| DL Model | Precision(%) | Recall(%) | F1(%) | Accuracy(%) |
| --- | --- | --- | --- | --- |
| ResNet | 97.83 ± 1.10 | 93.26 ± 2.02 | 95.49 ± 0.64 | 96.13 ± 0.50 |
| MobileNetV2 | 96.73 ± 1.71 | 95.13 ± 3.19 | 95.92 ± 1.65 | 96.44 ± 0.82 |

Both deep learning architectures achieved a comparable accuracy rate of 96%, effectively identifying the presence of blood vessels (Table 2). MobileNetV2 exhibited a precision of 97% and a recall of 95%. ResNet50, while having a slightly higher precision at 98%, had a lower recall of 93%. Despite the comparable accuracy, there was a slight difference in their F1 scores: MobileNetV2 maintained an F1 score of 96%, while ResNet50 had a slightly lower F1 score of 95%. The standard deviation values for precision, recall, F1 score, and accuracy are provided in Table 2, reflecting the performance consistency across different folds.

The confusion matrix for MobileNetV2 (Figure 3-a) demonstrates the model’s ability to accurately classify vessel and non-vessel regions. It revealed a high number of true positives for vessel detection and a low number of false negatives, showcasing its effectiveness. In contrast, non-vessel regions showed significant true negatives and minimal false positives. The probability distribution histogram (Figure 3-b) further illustrates MobileNetV2’s classification confidence, with two distinct peaks representing vessel and non-vessel images.

**Figure 3:**
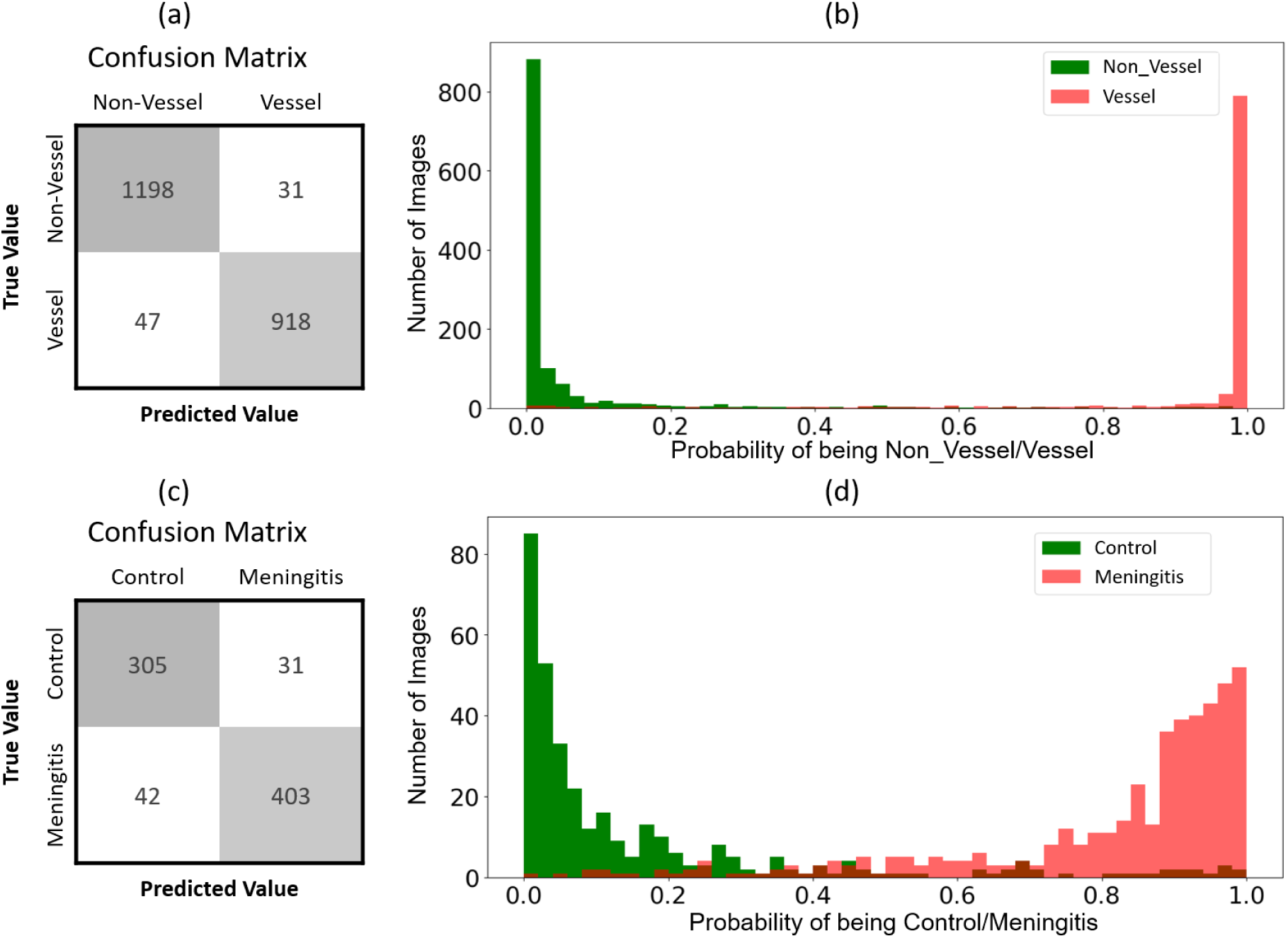
Results (a) Confusion Matrix of MobileNetV2 for Vessel and Non-Vessel Image Classification. (b) Probability Distribution Histogram of MobileNetV2 Classifications for Vessel and Non-Vessel Images: Histogram showing the probability of each image being classified as Healthy or Meningitis. (c) Confusion matrix of the ResNet50 model for Control or Meningitis classification. (d) Histogram showing the probability of each image being classified as control or meningitis.

### 3.2 Stage 2: Meningitis Screening at Patient Level

Stage 2 focuses on screening for *meningitis* cases. This stage uses a fine-tuned ResNet50 [12] architecture for binary classification at the image level. Next, we applied an ensemble technique to aggregate all the image scores corresponding to each patient into a final patient-level prediction.

At the image level, the ResNet50 model demonstrated a precision of 93%, recall of 90%, an F1 score of 92%, and an overall accuracy of 92%. The confusion matrix analysis revealed robust performance, with 305 true positives for control cases and 403 true positives for meningitis cases. The confusion matrix for meningitis vs control task is shown in Figure 3-c. The probability distribution for images being predicted as *meningitis* and *controls* shows a skewed U shape showing that there is a robust predictive power at the image level (Figure 3-d).

A key aspect of Stage 2 was the evaluation of ensemble learning techniques, particularly soft-voting and hard-voting methods [31], to aggregate image-level predictions for patient-level screening accuracy. Simultaneously, as illustrated in Table 3, our experiments examined the impact of different image sizes on this process. Lower resolution images offer advantages such as storage efficiency, faster handling, and reduced power consumption in devices. To achieve this, we employed column and row subsampling [32] to reduce the original image size of 556×200 pixels by factors of 2 and 4, resulting in experimental image sizes of 278×200, 139×200, 278×100, and 139×100 pixels. Our findings revealed that soft voting, which averages predicted probabilities, was more effective than hard voting across these downsized images, notably at smaller dimensions like 139×100 pixels, where accuracy markedly improved. Employing a fixed aggregation value of N = 7 predicted image outputs per patient, soft voting consistently outperformed hard voting, enhancing patient-level accuracy. This methodology successfully identified 15 out of 17 patient-level acquisitions at the smallest image size, demonstrating significant progress in precise patient-specific screening. Overall, the patient-level accuracy provided by this workflow reached 94%.

**Table 3.** Experiments Comparing Soft Voting and Hard Voting Techniques Across Various Image Sizes, “PLAccC” denotes the number of patients correctly classified out of a total of 17 patient-level acquisitions (16 patients, one with 2 LPs).

| Exp No. | Image Size | Single image |  |  | Soft Voting |  |  | Hard Voting |  |  |
| --- | --- | --- | --- | --- | --- | --- | --- | --- | --- | --- |
|  |  | Accuracy | Recall | PLAccC | Accuracy | Recall | PLAccC | Accuracy | Recall | PLAccC |
| 1 | 556x200 | 92.06 | 89.66 | 16 | 93.33 | 91.18 | 16 | 93.33 | 89.71 | 16 |
| 2 | 278x200 | 87.07 | 83.82 | 16 | 90.00 | 88.24 | 16 | 89.17 | 85.29 | 16 |
| 3 | 139x200 | 75.42 | 69.21 | 13 | 76.67 | 72.06 | 13 | 75.00 | 70.59 | 13 |
| 4 | 278x100 | 72.60 | 66.52 | 15 | 75.83 | 69.12 | 15 | 76.67 | 70.59 | 15 |
| 5 | 139x100 | 72.47 | 66.29 | 14 | 75.83 | 67.65 | 15 | 75.83 | 66.18 | 14 |

Next, it is relevant to understand the optimal number of N images needed per patient, as it determines the acquisition burden on the patient, the doctor, and the caregiver, as well as impacts the data storage and data processing times. Therefore, we conducted experiments with different numbers of images per participant (N = 3, 5, 7, and 9) to evaluate this aspect. As shown in Table 4, for the smallest image size (139×100 pixels), both soft and hard voting techniques were compared across various N values. This experiment was repeated 20 times with shuffled data, and Ttable 4 presents the mean results with standard deviation. Unsurprisingly, we found that performance in both image and patient-level accuracy improves as the number of images per patient N increases. However, soft voting effectively allows the information from all the images to be integrated providing an accurate result already at N = 7, allowing the system to make an effective compromise between accuracy and patient burden.

**Table 4.** Comparison of Soft and Hard Voting Based on aggregation values between 3-9 for the Smallest Image Size (139×100 Pixels). “PLAcc%” represents the patient level accuracy in percentage.

| N for Voting | Soft Voting |  |  | Hard Voting |  |  |
| --- | --- | --- | --- | --- | --- | --- |
|  | Accuracy | Recall | PLAcc% | Accuracy | Recall | PLAcc% |
| 3 | 77.74 ± 0.01 | 70.37 ± 0.01 | 83.82 ± 0.04 | 76.62 ± 0.01 | 69.00 ± 0.02 | 82.94 ± 0.04 |
| 5 | 78.61 ± 0.02 | 71.48 ± 0.02 | 82.06 ± 0.05 | 78.33 ± 0.01 | 70.49 ± 0.03 | 82.35 ± 0.03 |
| 7 | **79.13 ± 0.02** | **72.35 ± 0.04** | **87.18 ± 0.03** | **78.54 ± 0.01** | **70.88 ± 0.02** | **86.72 ± 0.03** |
| 9 | 79.95 ± 0.03 | 72.94 ± 0.04 | 84.71 ± 0.04 | 79.29 ± 0.02 | 71.57 ± 0.03 | 81.47 ± 0.02 |

### 3.3 Stage 3: Explainable AI (XAI)

In our study, we use Gradient-weighted Class Activation Mapping (GradCAM) [27], an XAI technique, to enhance the interpretability of our deep learning framework for the Quality Control and meningitis screening stages.

Grad-CAM, is a technique used for visualizing and understanding the decision-making process of deep neural networks, particularly convolutional neural networks (CNNs), in the context of image classification tasks, contributing to the explainability of an AI algorithm (XAI). It helps to highlight the regions of an input image that contribute the most to the final prediction made by the neural network. In a nutshell, GradCAM uses the gradient information flowing back from the final prediction to highlight the important regions in the input image. By visualizing these heatmaps, one can gain insights into which parts of the input image the model is paying attention to during the classification process.

**Quality Control XAI (Stage 1).** In the first stage, the deep learning classification model focused on distinguishing between *vessel* and *non-vessel* regions. GradCAM visualizations (Figure 4-a) highlighted the model’s strategy in differentiating these regions. Vessel regions, marked by red areas in the heatmap, showed structured high-frequency patterns, similar to barcodes. Conversely, non-vessel regions, which lacked such distinct patterns, demonstrated the model’s ability to identify areas without vessel presence.

**Figure 4.**
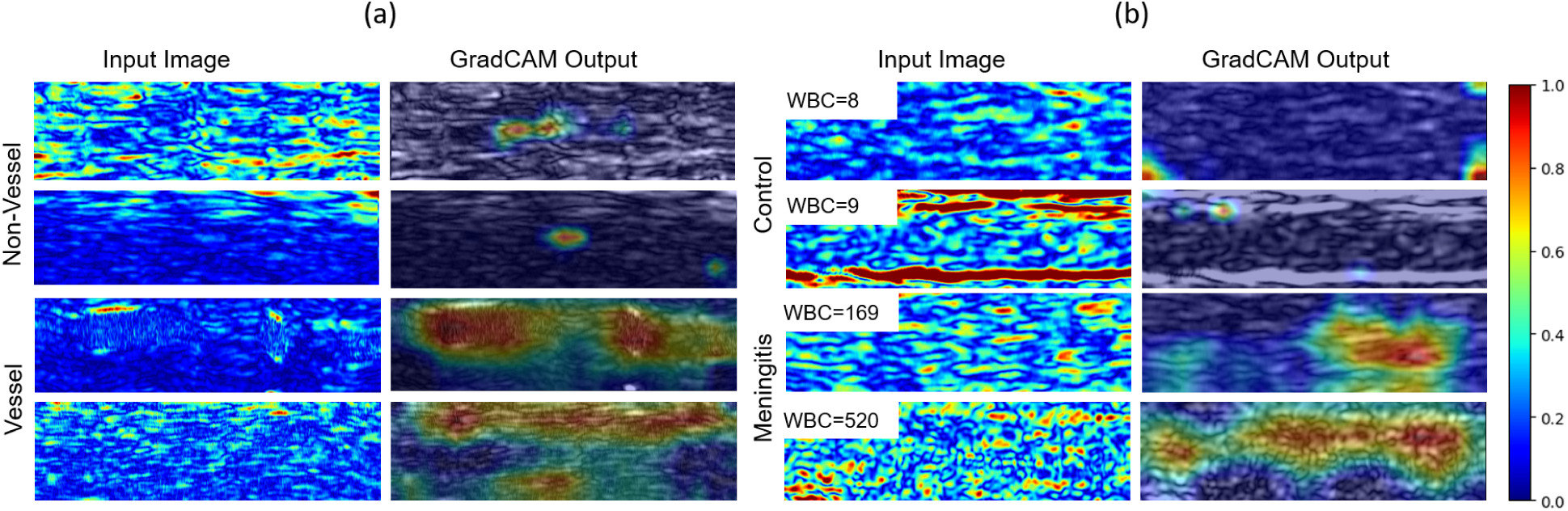
GradCAM heatmap visualizations: (a) Showcase patterns between vessel and non-vessel regions for Stage 1 classifications. (b) Features highlighted in control and meningitis cases, with WBC counts written in the top left corner. The accompanying color code bar represents the intensity scale of the heatmap, with red denoting regions of highest influence and blue indicating areas of least influence on the model’s predictive decision (same scale used for both vessel vs non-vessel and control vs meningitis models).

**Screening XAI (Stage 2).** In the second stage, aimed at distinguishing between *control* and *meningitis* cases, we also applied GradCAM to obtain a better insight into the model’s decision process. Initial examination of the HR ultrasound images (Figure 4-b) from both categories did not reveal distinguishable patterns to the human eye. Nevertheless, GradCAM visualizations revealed distinctive areas within the images, suggesting potential regions of interest associated with the presence of white blood cells (WBC), crucial indicators of meningitis. The heatmaps highlighted large, blob-like regions, resembling clusters, indicating the model’s areas of concentration without specifying diagnostic features.

To delve further into these observations, a statistics based XAI approach was utilized. Analysis of histograms for various image metrics, including normalized intensity, normalized intensity standard deviation, entropy, and the gradient in the Y direction, were calculated as shown in Figure 5-a. Distributions of all these metrics were checked using the JSD score to find differences between control and meningitis cases. Normalized pixel intensity distributions range between −1 and 1, showing a small peak at 1 indicative of image saturation (Figure 5-a top-left). Normalized intensity is higher for meningitis cases, indicative of higher image to noise ratio in cases vs controls. Notably, images from meningitis cases displayed a broader distribution in the intensity standard deviation and higher entropy values. This suggests a variation in complexity and variability within these images compared to those from control cases, which exhibited narrower and steeper distributions. Quantitatively, among all these metrics, entropy has the highest JSD score, indicating a notable distinction in this measure between *meningitis* and *control* categories.

**Figure 5.**
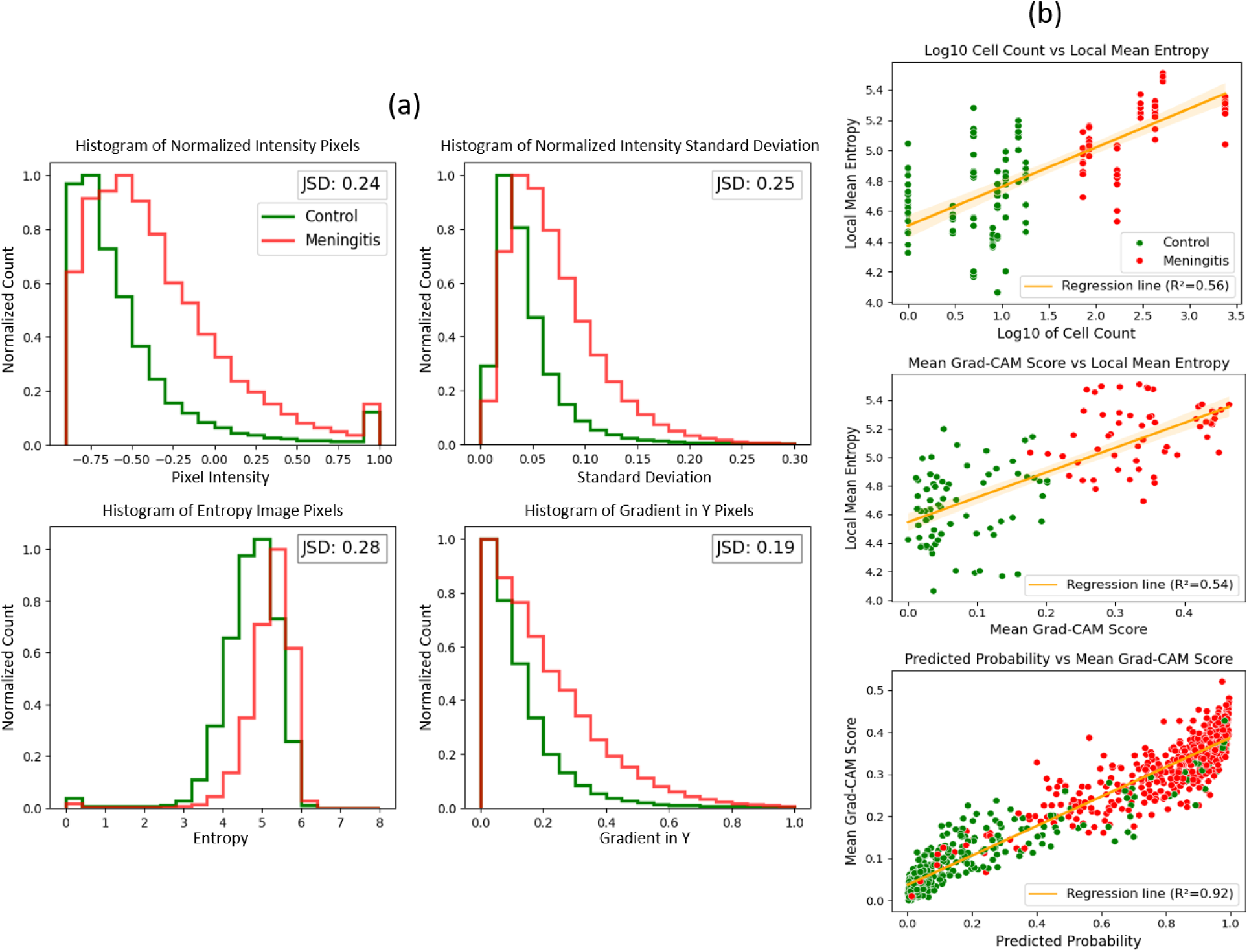
Statistical XAI analysis for meningitis screening: (a) Histograms ordered from top to bottom and left to right, comparing pixel intensity, normalized intensity standard deviation, entropy, and gradient in the Y direction between control and meningitis cases, with Jensen-Shannon Divergence (JSD) values quantifying the differences between distributions of meningitis cases and control images. Normalized pixel intensity values range between −1 to 1. (b) Scatter plots showing the relationship between cell count and local mean entropy (top), mean Grad-CAM scores and local mean entropy (middle), and predicted probabilities versus mean Grad-CAM scores (bottom), each with a regression line and R² value demonstrating the strength of correlation.

To further examine the nature of these “blobs” and their relationship to the white blood cells, we calculated the correlation between entropy versus cell count (top) and entropy versus mean Grad-CAM scores (Figure 5-b, middle). The analysis revealed a moderate correlation for both, with R² values of 0.56 for entropy and cell count, and 0.54 for entropy and mean Grad-CAM scores. This observation suggests a proportional relationship between the complexity captured by entropy in the ultrasound images and the presence of WBCs.

The bottom plot of Figure 5-b illustrates the correlation between mean Grad-CAM scores and predicted probabilities for each image. Figure 5-b (bottom) shows a very good separation between cases and controls, accounting for the high performance of the classifier. Control cases predominantly clustered in the lower left quadrant, showing lower Grad-CAM scores and predicted probabilities, whereas meningitis cases were mainly found in the upper right quadrant, associated with higher scores and probabilities. The R² value of 0.92 indicates a significant correlation, illustrating the relationship between the model’s Grad-CAM activations and the predicted likelihood of meningitis.

## 4. Discussion

Our study introduces a multistage deep learning approach for non-invasive screening of infant meningitis, a life-threatening condition, utilizing HR ultrasound images acquired below the infant fontanelle region. This method addresses, in a first step, many of the challenges associated with lumbar puncture (LP), the established gold standard for meningitis diagnosis. LP, an invasive procedure involving spinal fluid extraction, is not innocuous and can be particularly distressing for infants and their families. Moreover, the requirement for skilled execution and specialized equipment and laboratory infrastructures makes LP and its subsequent analysis challenging in resource-limited settings, where such medical resources are often scarce. Our approach is designed to potentially limit the use of LP to only high-suspicion and positive cases by providing preliminary noninvasive screening based on WBC count in CSF through an AI-based automated ultrasound framework, designed for real-time application in clinical settings. Ultimately, accurate discrimination of WBC levels in CSF could significantly narrow down indications of LPs, sparing unnecessary LPs to the majority of suspected cases that are ultimately ruled out, and also saving costs and valuable resources, both in high- and low-income countries. Conversely, a high suspicion of meningitis, as suggested by a high WBC count determined by the initial screening, would help target LPs in those cases whereas the likelihood of the diagnosis is very high.

The first stage of the pipeline dedicated to image quality control, focusing on the identification of blood vessels with 2194 ultrasound images, achieved a solid accuracy of 96% using the MobileNetV2 architecture. This accuracy is critical, as the classification process is embedded directly into the diagnostic devices, enabling clinicians to conduct real-time screening and ensuring that only high-quality images, free from blood vessels, are captured. This approach optimizes storage and processing time by skipping the acquisition of images that don’t meet quality standards. MobileNetV2 has a simpler architecture, with only 3.5 million trainable parameters compared to ResNet50’s 25.6 million. This simplicity aligns with the real-time hardware requirements of clinical settings, which demand quick processing and efficiency. Accurate, automatic, and on-the-fly image quality control methods are key diagnostic and screening platform components. Our future developments in quality control involve extending beyond vessel identification to fully automated filters that include clutter, poor quality, and other artifacts in ultrasound scans.

Stage 2, the patient-level screening phase, employed a fine-tuned ResNet50 model on a refined dataset of 781 images from 16 patients. This stage differentiated between *control* (<30 WBC/mm³) and *meningitis* cases (≥30 WBC/mm³). We achieved a precision of 93% and an accuracy of 92% in distinguishing between *control* and *meningitis* instances based on imaging frames. Notably, at the patient level, the stage accurately identified 6 *meningitis* cases and 10 negative *controls* among 16 patients (one patient with two LPs, resulting in 17 LPs), achieving a sensitivity of 100% and a specificity of 90%, with only a single *control* misclassification.

Moreover, our study highlighted the crucial role of ensemble learning techniques, particularly soft voting, in enhancing diagnostic accuracy at the patient level. Soft voting, by averaging predicted probabilities, proved superior to hard voting, especially when analyzing images at reduced resolutions. For instance, in the smallest image dimension of 139×100 pixels, soft voting not only improved accuracy from 72% to 78% but also correctly reclassified an initially misdiagnosed patient. This approach enabled more effective patient-level screening, demonstrating the model’s robust predictive power even with downsized images. By employing column and row subsampling, we not only optimized storage and processing efficiency but potentially minimized ultrasound scan time exposure, aligning with our goal to reduce patient discomfort and streamline the screening workflow.

The choice of N=7 for aggregating consecutive image frame results was found optimal, striking a balance between accuracy and processing demand. While increasing the number of frames does enhance accuracy by leveraging the predicted probabilities per image, the performance gains start to saturate, presenting diminishing returns. This consideration is crucial in practical settings, particularly in real-time clinical scenarios where processing time is a critical factor. Additionally, a shorter acquisition time implies less burden to the patient and less time required by the clinician, further emphasizing the significance of this optimal frame aggregation choice.

In stage 3, the integration of Explainable AI (XAI) techniques, particularly Gradient-weighted Class Activation Mapping (GradCAM), provides insights into our model’s decision-making processes, enhancing transparency and trust in our AI-driven approach. This approach in the initial quality control phase highlighted that the model identifies distinct high-frequency patterns in vessel images, similar to barcodes, which are visible to the human eye. These patterns, indicating the presence of vascular structures, are key for the model’s ability to distinguish between *vessel* and *non-vessel* images. This observation indicates that the model mimics a human-like visual assessment process, relying on visible patterns to classify images accurately.

XAI proves particularly useful in Stage 2, where structural differences between control and meningitis cases are not easily visible to the human eye. The backscatter signals from potentially present WBCs in the CSF generate a pattern that is not always easily interpretable and distinguishable from clean CSF by visual inspection. As illustrated in Figure 4-b, the GradCAM heat map identifies larger areas or “blobs’’ in the meningitis case images. This observation suggests that the network learns from the long-range patterns created by the complex interaction between the ultrasound waves and the white blood cells, using these areas as clues to differentiate between the two classes.

To understand these differences better, we applied a statistical-based XAI approach to complement GradCAM. This method involved analyzing intensity variations, entropy, standard deviation, and gradients to quantify the variances hinted at by GradCAM visualizations. We selected these metrics based on our hypothesis that the presence of WBCs, indicative of meningitis, would result in quantifiable statistical changes in the ultrasound images. These changes would become evident when comparing *control* to *meningitis* images. The results, presented in Figure 5-a, confirmed our hypothesis. We observed that meningitis cases had higher average values for entropy and standard deviation, reflecting increased image complexity and variability which are key factors in distinguishing between *meningitis* and *control* images. The Jensen-Shannon Divergence (JSD) values further quantified the differences between the distributions of these metrics for the two groups. These higher values are indicative of the textural influences of backscatter signals from WBCs present in the CSF in the meningitis images, which our model relies on to differentiate between conditions.

The JSD score for the entropy histogram, at 0.28, is the highest difference among all evaluated metrics, effectively distinguishing *meningitis* cases from *control* images. Entropy is a statistical measure that quantifies the information content within images, indicating that images with greater variability and complexity, characteristics indicative of meningitis, tend to exhibit higher entropy. This suggests that images with higher entropy levels are characterized by significant variations, shown as complex and irregular patterns in pixel intensity due to the presence of both isolated or clustered white blood cell (WBC) dispersions or clusters. It needs to be noted, however, that while the entropy is significantly higher in images of meningitis patients, there is still a large local mean entropy variability in control images which we attribute to several sources of noise including clutter. For that reason, entropy and intensity measures are insufficient for classifying whether an image originates from a meningitis or control patient. The advantage of the deep learning classifier resides in its ability to learn complex patterns beyond texture, intensity and gradient, learning to differentiate signal from noise, and thereby providing an accurate classification performance.

Moreover, as shown in the scatter plot in Figure 5-b, there is a positive correlation between the cell count and the local mean entropy, evidenced by the regression line with an R² value of 0.56. This correlation suggests that as WBC counts rise, as an increasing sign of infection, so does the entropy in the image, suggesting a greater level of complexity due to WBC aggregation. Similarly, the average Grad-CAM scores, which identify the regions most significant for diagnosis according to the neural network, also have a positive correlation with mean entropy, as shown by a regression line with an R² of 0.54. This indicates that the model focuses on areas with higher entropy, likely reflecting a greater presence of WBCs, for its classification decisions. Taken together, these observations indicate that the neural network can not only detect the presence but also the quantity of white blood cells. This raises the prospect of developing more sophisticated multi-class or regression-based models for future studies with larger datasets.

The comprehensive application of XAI, merging GradCAM visualizations with statistical analysis, enhances our understanding of the diagnostic features within ultrasound images, facilitating a deeper insight into the model’s decision-making process in distinguishing meningitis from control cases.

Our study has limitations. One main constraint for deep learning model training is the relatively small size of our patient cohort, comprising only 17 acquisitions from 16 individuals with a total of 781 images for Stage 2 analysis. To address this challenge, we opted to use a leave-one-out cross-validation strategy, allowing us to assess each patient’s data individually and potentially enhance the robustness of our findings. With this strategy, the model’s accuracy could still be robustly evaluated on an unseen sample of patient images. Future validation of prospective datasets from a diverse set of patients is needed for the implementation of a production algorithm, and patients are already being recruited in Spain, Mozambique, and Morocco for that purpose.

Another limitation of our study is the use of a threshold of 30 cells/mm³ to differentiate between *meningitis* and *control* cases. While this threshold has been effective in classifying patients within our cohort, which includes children from various age groups, its applicability might not be universal across different hospitals, age groups, and disease stages. The factor of age is especially important, as the thresholds for white blood cell counts vary with age, highlighting the necessity of considering age when diagnosing meningitis to ensure accuracy across a broad spectrum of patient ages. Also, in specific scenarios (such as immunocompromised patients, some viral meningitis or CSF exams after receiving antibiotics), meningitis can be present without an elevated WBC count in CSF. However, the aim of the device is to be equal to the laboratory WBC count in CSF, therefore in such gray area scenarios, clinicians should interpret Neosonics’ results as they would the gold standard and decide accordingly. For instance, if the clinical suspicion is high independently of the WBC count in CSF, LPs should be performed for further microbiological investigation. On the other hand, Neosonics could be performed in scenarios where now clinicians face great uncertainty and are forced to make decisions without certainty. One example is very severe and unstable patients in which LPs are contraindicated and, therefore, delayed, and clinicians need to treat them empirically. Also, some patients present with slightly elevated WBC counts in CSF but are not considered meningitis cases and, accordingly, are not treated for it, and clinicians could easily repeat non-invasive exams periodically to ensure that inflammatory cells go back to normal values without treatment.

Additionally, the process of cell counting with a Fuchs-Rosenthal or Neubauer chamber following lumbar puncture (LP) involves technicians analyzing cells across 8 out of 64 quadrants under a microscope and taking the average to determine the count for each patient. While this method is the gold standard, it is subject to human error and variability introduced by the technicians, thereby affecting the precision of the ground truth data. For instance, the visualization of a single cell in the 8/64 count can make final results differ by 8 cells, which can be even higher if the initial sample has been diluted prior to its analysis (due to high cellularity, either in WBC or blood contamination during the procedure) and, consequently, conversion factors need to be applied to obtain the final result. This problem is even more exacerbated in LICs, where the baseline resources and technicians’ training are poorer. This partially explains the variability observed in the graph of cell count vs. entropy graph in Figure 5-b (top) of our study.

As part of our future steps, we aim to provide clinicians with quantitative results, offering high sensitivity even at low cell concentrations (e.g., 5 cells/mm³). By incorporating a robust regression or multi-classification model, we plan to develop a refined method that adjusts for inherent errors and improves accuracy across a wider range of cell concentrations. This quantitative approach will enable clinicians to interpret results based on their specific thresholds, which may vary according to clinical context (such as age, previous antibiotics administration or immunocompromised patients).

The clinical impact of our study is considerable, particularly given that most patients in our cohort were subjected to lumbar punctures (LPs) because of non-specific symptoms like fever and raised inflammatory markers. By employing deep learning (DL) models with our non-invasive ultrasound method to estimate the WBC count in the CSF, we offer a promising approach for meningitis screening in neonates and infants. This could notably decrease the necessity for LPs, facilitating more precise indications for LPs, quicker commencement of treatment, and improved outcomes for patients.

In summary, our research marks a significant advancement in the use of AI and deep learning for the non-invasive screening of meningitis in neonates and infants. By integrating advanced AI models with ultrasound imaging, we’ve introduced a novel and non-invasive approach that has the potential to transform the preliminary screening process for meningitis. This approach not only has the potential to substantially decrease the reliance on unnecessary lumbar punctures but also highlights the profound impact of AI in democratizing health care by facilitating efficient and affordable screening technology to areas with high need, where there is limited availability of laboratory resources. Moreover, our work highlights the importance of explainability and transparency in AI required for all medical applications where the objective is clinical decision support to screening and diagnosis. Our utilization of eXplainable AI (XAI) has demonstrated that increases in complexity within ultrasound imaging, such as gradient changes, which may go unnoticed by the human eye, are correlated with higher blood cell counts and thus indicative of greater disease severity.

Moving forward, our goal is to further refine these AI-driven methodologies to ensure their effectiveness and applicability in a wider range of clinical scenarios. The progress outlined in our study opens new pathways in neonatal and infant healthcare, where AI enhanced technologies provide safer, more accessible, and efficient diagnostic options, establishing a new benchmark for the early detection and management of life-threatening conditions like meningitis.

## Data Availability

All data produced in the present study are available upon reasonable request to the authors

## Ethical Approval

The Ethics Committee of Clinical investigation of the University Hospital La Paz (Madrid) gave ethical approval for this work (Approval reference number 5183).

## Acknowledgments

ISGlobal acknowledges support from the grant CEX2018-000806-S funded by MCIN/AEI/ 10.13039/501100011033, support from the Generalitat de Catalunya through the CERCA Program, support from the Ministry of Research and Universities of the Government of Catalonia (2021 SGR 01563), support from the grant INV-048197 funded by the Bill and Melinda Gates Foundation, and from Instituto de Salud Carlos III (FIS PI16/00738). Kriba acknowledges support from the European Union’s Horizon Europe research and innovation programme under project code 190155553 - NEOSONICS. CISM is supported by the Government of Mozambique and the Spanish Agency for International Development (AECID).

We acknowledge the contributions of the UNITED Study Consortium members: Manuela Lopez-Azorín, Eva Valverde, Marta Ybarra, M. Carmen Bravo, Carles Luaces, David Muñoz, Thais Agut, Barbara Salas, Nuria Carreras, Ana Alarcón, Martín Iriondo, Alberto Ibáñez, Montserrat Parrilla, Luis Elvira, Cristina Calvo, Adelina Pellicer, Fernando Cabañas, Justina Bramugy, Muhammad Sidat, Mastalina Zandamela, Paula Rodrigues, Dulce Graça, Sebastião Ngovene, Anelsio Cossa, Janeta Machai, Campos Mucasse, Luzidina Martins and Sandra Massango for their instrumental efforts in the completion of this work

## Declaration of Generative AI and AI-Assisted Technologies in the Writing Process

During the preparation of this work, the authors used ChatGPT and DeepL to improve the readability of the manuscript and as a translation aid since some co-authors do not have English as their native language. After using these tools, the authors reviewed and edited the content as needed and take full responsibility for the content of the published article.

## References

1. Schiess N, Groce NE, Dua T. The Impact and Burden of Neurological Sequelae Following Bacterial Meningitis: A Narrative Review. Microorganisms 2021; 9(5)

2. Global, regional, and national burden of meningitis and its aetiologies, 1990-2019: a systematic analysis for the Global Burden of Disease Study 2019. Lancet Neurol 2023; 22(8): 685–711.

3. Glatstein MM, Zucker-Toledano M, Arik A, Scolnik D, Oren A, Reif S. Incidence of traumatic lumbar puncture: experience of a large, tertiary care pediatric hospital. Clin Pediatr (Phila) 2011; 50(11): 1005–9.

4. Dalai R, Dutta S, Pal A, Sundaram V, Jayashree M. Is Lumbar Puncture Avoidable in Low-Risk Neonates with Suspected Sepsis? Am J Perinatol 2022; 39(1): 99–105.

5. Flidel-Rimon O, Leibovitz E, Eventov Friedman S, Juster-Reicher A, Shinwell ES. Is lumbar puncture (LP) required in every workup for suspected late-onset sepsis in neonates? Acta Paediatr 2011; 100(2): 303–4.

6. Bedetti L, Marrozzini L, Baraldi A, et al. Pitfalls in the diagnosis of meningitis in neonates and young infants: the role of lumbar puncture. J Matern Fetal Neonatal Med 2019; 32(23): 4029–35.

7. Lee, June-Goo, et al. “Deep learning in medical imaging: general overview.” Korean journal of radiology 18.4 (2017): 570–58

8. Krizhevsky, Alex, Ilya Sutskever, and Geoffrey E. Hinton. “Imagenet classification with deep convolutional neural networks.” Advances in neural information processing systems 25 (2012).

9. Simonyan, Karen, and Andrew Zisserman. “Very deep convolutional networks for large-scale image recognition.” arXiv preprint arXiv:1409.1556 (2014).

10. Szegedy, Christian, et al. “Going deeper with convolutions.” Proceedings of the IEEE conference on computer vision and pattern recognition. 2015.

11. Howard, Andrew G., et al. “Mobilenets: Efficient convolutional neural networks for mobile vision applications.” arXiv preprint arXiv:1704.04861 (2017).

12. He, Kaiming, et al. “Deep residual learning for image recognition.” Proceedings of the IEEE conference on computer vision and pattern recognition. 2016.

13. Lakhani P, Sundaram B. deep learning at chest radiography: automated classification of pulmonary tuberculosis by using convolutional neural networks. Radiology. 2017;284:574–82.

14. Gulshan V, Peng L, Coram M, et al. Development and validation of a deep learning algorithm for detection of diabetic retinopathy in retinal fundus photographs. JAMA. 2016;316:2402–10.

15. Esteva A, Kuprel B, Novoa RA, et al. Dermatologist-level classification of skin cancer with deep neural networks. Nature. 2017;542:115–8.

16. Shorten, Connor, Taghi M. Khoshgoftaar, and Borko Furht. “Deep Learning applications for COVID-19.” Journal of big Data 8.1 (2021): 1–54.

17. Debelee, Taye Girma, et al. “Survey of deep learning in breast cancer image analysis.” Evolving Systems 11 (2020): 143–163.

18. N. Buda, E. Segura-Grau, J. Cylwik, and M. Wełnicki, “Lung ultrasound in the diagnosis of COVID-19 infection - a case series and review of the literature,” Advances in Medical Sciences, vol. 65, no. 2, pp. 378–385, 2020.

19. G. Soldati, A. Smargiassi, R. Inchingolo et al., “Is there a role for lung ultrasound during the COVID-19 pandemic?,” Journal of Ultrasound in Medicine, vol. 39, no. 7, pp. 1459–1462

20. Meng, Dan, et al. “Liver fibrosis classification based on transfer learning and FCNet for ultrasound images.” Ieee Access 5 (2017): 5804–5810.

21. Liu, Xiang, et al. “Learning to diagnose cirrhosis with liver capsule guided ultrasound image classification.” Sensors 17.1 (2017): 149.

22. Chi, Jianning, et al. “Thyroid nodule classification in ultrasound images by fine-tuning deep convolutional neural network.” Journal of digital imaging 30 (2017): 477–486.

23. Jimenez, Xavier, et al. “Quantification of very low concentrations of leukocyte suspensions in vitro by high-frequency ultrasound.” Ultrasound in Medicine & Biology 42.7 (2016): 1568–1573.

24. Lee, John H., Duane S. Boning, and Brian W. Anthony. “Measuring the absolute concentration of microparticles in suspension using high-frequency B-mode ultrasound imaging.” Ultrasound in Medicine & Biology 44.5 (2018): 1086–1099.

25. Elvira, Luis, et al. “A New Methodology for the Assessment of Very Low Concentrations of Cells in Serous Body Fluids Based on the Count of Ultrasound Echoes Backscattered From Cells.” IEEE Transactions on Ultrasonics, Ferroelectrics, and Frequency Control 68.5 (2020): 1580–1592.

26. Fernandez, Alba, et al. “Estimation of the concentration of particles in suspension based on envelope statistics of ultrasound backscattering.” Ultrasonics 116 (2021): 106501.

27. Selvaraju, Ramprasaath R., et al. “Grad-cam: Visual explanations from deep networks via gradient-based localization.” Proceedings of the IEEE international conference on computer vision. 2017.

28. Hastie, Trevor, et al. The elements of statistical learning: data mining, inference, and prediction. Vol. 2. New York: springer, 2009.

29. Kingma, Diederik P., and Jimmy Ba. “Adam: A method for stochastic optimization.” arXiv preprint arXiv:1412.6980 (2014).

30. Kiefer, Jack, and Jacob Wolfowitz. “Stochastic estimation of the maximum of a regression function.” The Annals of Mathematical Statistics (1952): 462–466.

31. Rokach, Lior. “Ensemble-based classifiers.” Artificial intelligence review 33 (2010): 1–39.

32. Gonzalez, Rafael C., and Richard E. Woods. Digital Image Processing. 4th ed., Pearson, 2018.

33. Arrieta, Alejandro Barredo, et al. “Explainable Artificial Intelligence (XAI): Concepts, taxonomies, opportunities and challenges toward responsible AI.” Information fusion 58 (2020): 82–115.

34. Amann, Julia, et al. “Explainability for artificial intelligence in healthcare: a multidisciplinary perspective.” BMC medical informatics and decision making 20 (2020): 1–9.

35. Cinà, Giovanni, et al. “Why we do need explainable ai for healthcare.” arXiv preprint arXiv:2206.15363 (2022).

36. Srinivasu, Parvathaneni Naga, et al. “From blackbox to explainable AI in healthcare: existing tools and case studies.” Mobile Information Systems 2022 (2022): 1–20.

37. Ajanovic, Sara, et al. “Meningitis Screening in Young Infants Based on a Novel, Non-Invasive, Transfontanellar Ultrasound Device: a Proof-of-Concept Study.” (2023). PREPRINT available at [10.21203/rs.3.rs-3677475/v1].

38. Song, Di, et al. “A new xAI framework with feature explainability for tumors decision-making in Ultrasound data: comparing with Grad-CAM.” Computer Methods and Programs in Biomedicine 235 (2023): 107527.

39. Nguyen, Truong Thanh Hung, et al. “Towards Trust of Explainable AI in Thyroid Nodule Diagnosis.” arXiv preprint arXiv:2303.04731 (2023).

40. Hasan, Md Mahmodul, et al. “FP-CNN: Fuzzy pooling-based convolutional neural network for lung ultrasound image classification with explainable AI.” Computers in Biology and Medicine 165 (2023): 107407.

41. McKinney, Wes. “pandas: A Foundational Python Library for Data Analysis and Statistics.” Python for High Performance and Scientific Computing, 14 Nov. 2010.

42. Yang, Wenzhuo, et al. “OmniXAI: A Library for Explainable AI.” 2022, arXiv:2206.01612

43. scikit-image: Image processing in Python.” Version 0.18.3, SciPy community, 2021. https://scikit-image.org/.

44. Bradski, G. “The OpenCV Library.” Dr. Dobb’s Journal of Software Tools, 2000.

45. Cover, Thomas M., and Joy A. Thomas. Elements of Information Theory. 2nd ed., Wiley-Interscience, 2006.

46. Virtanen, Pauli, et al. “SciPy 1.0: fundamental algorithms for scientific computing in Python.” Nature methods 17.3 (2020): 261–272.

47. Chollet, François, et al. “Keras.” 2015, https://keras.io.

48. Pedregosa, Fabian, et al. “Scikit-learn: Machine learning in Python.” the Journal of machine Learning research 12 (2011): 2825–2830.

49. Aerts, Céline, et al. “Cost-Effectiveness Analysis of Implementing a Non-Invasive Screening Tool (Neosonics) for Meningitis Among Newborns in Mozambique, Morocco and Spain.” Meningitis Research Foundation Conference 2023

50. Jiménez, Javier, et al. ’System and Method for Non-invasive White Blood Cell Counting in Serous Body Fluids.’ Patent PCT/EP2023/064154. World Intellectual Property Organization, 26 Apr. 2023.

